# School-based programme to reduce dietary salt intake and blood pressure amongst adolescents and their parents: a cluster-randomised controlled trial in rural and urban Malawi

**DOI:** 10.64898/2025.12.22.25340861

**Authors:** Stefan Witek-McManus, Nozgechi Phiri, James R. Carpenter, Shekinah Munthali-Mkandawire, McDonald Chabwera, Miryam Katundulu, Caroline Chiphinga-Mwale, Rachel Burton, Marko Kerac, Emma McIntosh, Dalitso Dembo Kang’ombe, Albert Saka, Frances S. Mair, Judith R. Glynn, Amelia C. Crampin, Jones Masiye

## Abstract

1.

**Background:** Excess dietary salt intake and high blood pressure are growing public health challenges in Africa, with limited evidence for effective interventions. We evaluated the effect of a school-based programme on salt intake and blood pressure of adolescents and their parents.

**Methods:** We conducted a cluster-randomised controlled trial with 26 primary schools in rural and urban Malawi. All pupils aged 11–14 years and their parents were eligible. The intervention comprised lessons and activities delivered over one school term promoting reduced salt intake. Co-primary outcomes were change in adolescent salt intake and systolic blood pressure (SBP) at 3-months. Secondary outcomes were change in adolescent diastolic blood pressure (DBP), adult salt intake, and adult blood pressure at 3-months; and change in blood pressure of all participants at 12-months.

**Findings:** We recruited 1968 participants between September 24^th^–October 13^th^ 2019 and January 8^th^–24^th^ 2020. Thirteen primary schools received the salt reduction programme (362 adolescents, 614 adults) and 13 the routine curriculum (362 adolescents, 596 adults). At baseline, mean urinary salt was 3·9g/day (SD 2·15) for adolescents and 4·3g/day (SD 2·50) for adults. Participation in the intervention was high, with overall compliance of 87% by adolescents. At 3-months, there was no significant difference in change in adolescent salt intake (-0·17g/day, 95% CI -0·63–0·29) or adolescent SBP (0·59mmHg, -1·80–2·97). There was also no significant difference in change in adult salt intake, or blood pressure at 3 or 12 months, except adolescent SBP at 12-months.

**Interpretation:** A school-based salt reduction intervention was feasible and had high levels of engagement, but did not reduce salt intake or blood pressure of adolescents and adults over a 3-month period. Longer term implementation, complemented by broader strategy and policy engagement, may be needed for school-based approaches to achieve reductions in dietary salt and blood pressure.

**Funding:** Medical Research Council

**Registration:** ISRCTN13909759

**RESEARCH IN CONTEXT:** *Evidence before this study:*

- Despite evidence that population-level dietary salt (sodium chloride) intake in Africa exceeds global recommendations, rigorous evidence for effective public-health strategies to address this are lacking. School-based approaches are widely used in Africa for the delivery of preventive health services, and may present a practical and efficient platform for addressing excess dietary salt intake.
- A trial previously conducted in urban China demonstrated the feasibility of reducing salt intake amongst school-age children, but was conducted in a setting of relatively high excess salt intake and a better-resourced education system. Previous studies in Africa have demonstrated the feasibility and effectiveness of school-children as health ‘messengers’ within their household, but this approach has not been evaluated for non-communicable diseases.

*Added value of this study:*

- We report the first randomised controlled trial of a school-based salt reduction programme to be conducted in Africa; and the first in a setting that is low-income, rural, or of moderate excess salt intake. Study strengths include recruitment across both rural and urban settings, enhancing generalisability, and the use of 24-hour urine collection, providing robust assessment of salt consumption.
- At baseline, we observed lower estimates of dietary salt intake than previously estimated through spot urine samples, equating to 27% of child and 33% of adult participants consuming dietary salt in excess of WHO recommendations. After 3-months, the average salt intake of school-children or adults who received the intervention did not significantly reduce overall after intervention implementation for one school term, compared to those who continued to receive the routine school curriculum (control).

*Implications of all the available evidence:*

- Salt-reduction strategies that leverage cross-sectoral partnerships, engaging education, health, and community actors, present a pragmatic approach to addressing excess salt intake in Africa. Our results demonstrate the feasibility of implementing such interventions within schools to reach both children and adults.
- While we did not observe significant reductions in salt intake, this may reflect the short implementation period and moderate baseline salt intakes in our study population. School-based interventions may need to be sustained over multiple years to achieve meaningful reductions in dietary salt intake, combined with complementary strategies including community engagement, school meal policies, and food environment modifications.
- Future research should consider evaluation of broader school-based interventions that are implemented over extended periods, embedded within comprehensive population-level salt reduction strategies.

## 3. INTRODUCTION

Excess salt intake is associated with elevated blood pressure (BP) and hypertension; the predominant risk factor for cardiovascular disease (CVD) and the largest single contributor to the global burden of disease [1]. Consumption of salt in excess, defined by the World Health Organisation (WHO) as more than >5 grams/day for adults, is widely observed across a variety of settings globally and was estimated to cause 1.65 million annual deaths attributable to cardiovascular causes in 2010 [2]. In response to this challenge, a global target of 30% reduction in salt intake and 25% reduction in the prevalence of raised BP has been proposed, with salt-reduction recommended as a cost-effective ‘best buy’ in the control of non-communicable diseases (NCD) [3].

While many high-income settings have begun to see success over the past decade in population-level reductions in salt intake and concomitant reductions in BP, there is growing evidence that both salt intake [4], and the prevalence of hypertension in adults [5] continue to increase in Africa. In 2017, CVD represented the second largest cause of death, and the largest single contributor to the burden of NCDs within the region [6]. However, little progress has been made in implementing effective prevention strategies [7], with a dearth of evidence from Africa for interventions that are effective at reducing salt intake [8] or primary hypertension [9].

Leveraging the scale-up of universal primary education, school-health strategies are widely used in low-income settings to reach children with basic health interventions such as deworming and school-feeding programmes [10], and are increasingly used to tackle NCD risk factors in middle and high-income settings [11], including as a mechanism to promote health behaviours among parents and community members [12]. Public health strategies to address raised BP have also begun to advocate for approaches that begin in childhood in response to evidence that hypertension is increasingly common at younger ages, elevated BP early in life tracks into adulthood; and that dietary preferences are challenging to effect later in life [13].

Building on the established school-health platform and high primary school enrolment in Malawi, we conducted a cluster-randomised trial to test the hypothesis that a school-based programme which educated adolescents about excess salt, and supported them to promote salt reduction within their households, would reduce the salt intake and blood pressure of adolescents and their parents.

## 4. METHODS

### 4.1. Ethical considerations

The trial was approved by the National Health Science Research Commission of Malawi (NHSRC) (#2206) and London School of Hygiene & Tropical Medicine (LSHTM) Research Ethics Committee (#17098). All adolescents enrolled in standard 6 at intervention schools were eligible to participate in the intervention. Written consent for the study assessments with adolescents was sought from their parent or caregiver, in addition to written assent from adolescents, and written consent from all adult participants. All information and consent procedures were conducted in the participants’ local language (Chichewa or Chitumbuka). Participants identified as hypertensive (mean SBP≥140mmHg, mean DBP≥90mmHg, or taking antihypertensive medication) during the survey assessments were eligible to participate, but also referred to care at a local primary healthcare facility if not previously diagnosed.

### 4.2. Study design

The intervention was evaluated using a two-arm cluster-randomised controlled design in which 26 primary schools were equally randomised (1:1) to implementation of the school-based salt reduction programme over one school term (3 months) or routine school curriculum, whereby no intervention was implemented. Randomisation was stratified by location (Karonga/Lilongwe), district (rural/urban) and school term (Oct-Dec/Jan-Mar). The study timeline is illustrated in Supplementary Figure 1. Due to the nature of the intervention, participants and outcome assessors were not blinded. However, efforts to maintain blinding to the intervention focus of dietary salt were made by inclusion of other dietary components (sugar and fat/oils) as part of the study procedures, as recommended elsewhere [14].

The power calculation was adjusted for clustering (ICC 0·05) and estimated at the 2-sided 5% level. Assuming a control mean salt intake level of 5g/day (SD 2.5) and sample size of 20 adolescents in 26 school would give 79% power to detect a relative reduction of 17% (i.e. 0·85 g/day absolute) in salt intake, and >90% power to detect a 20% relative (1 g/day absolute) reduction in salt intake. Assuming two adults enrolled per adolescent (1040 adults) and the same ICC, type-1 error and control-mean salt level of 5g/d (SD 2.5), there would be 89% power to detect a 17% relative (i.e. 0·85 g/day absolute) reduction in salt intake, and 96% power to detect a 20% relative (1 g/day absolute) reduction in salt intake.

### 4.3. Study setting

The trial was conducted in two regions of Malawi (Figure 1A): Areas 25, 48 & 49 of Lilongwe district and Karonga HDSS (Health and Demographic Surveillance Site) in Karonga district (Figure 1B). Karonga HDSS comprises a predominantly rural setting located in the far north of Malawi, with an economy based predominantly on subsistence agriculture and fishing from Lake Malawi. In contrast, Areas 25, 48 & 49 of Lilongwe district are areas of relatively high population density located to the east and north-east of the city centre, with an economically mixed profile. Community-based surveys conducted in both sites between 2013-2016 reported 50-53% of participants to reside in households with an estimated per-capita salt intake of >5g/day based on household-level reporting of plain salt usage, and 91-96% of a sub-sample of adults had 24-h urinary sodium >2 g/day (by spot urine sample) [15].

**Figure 1A:**
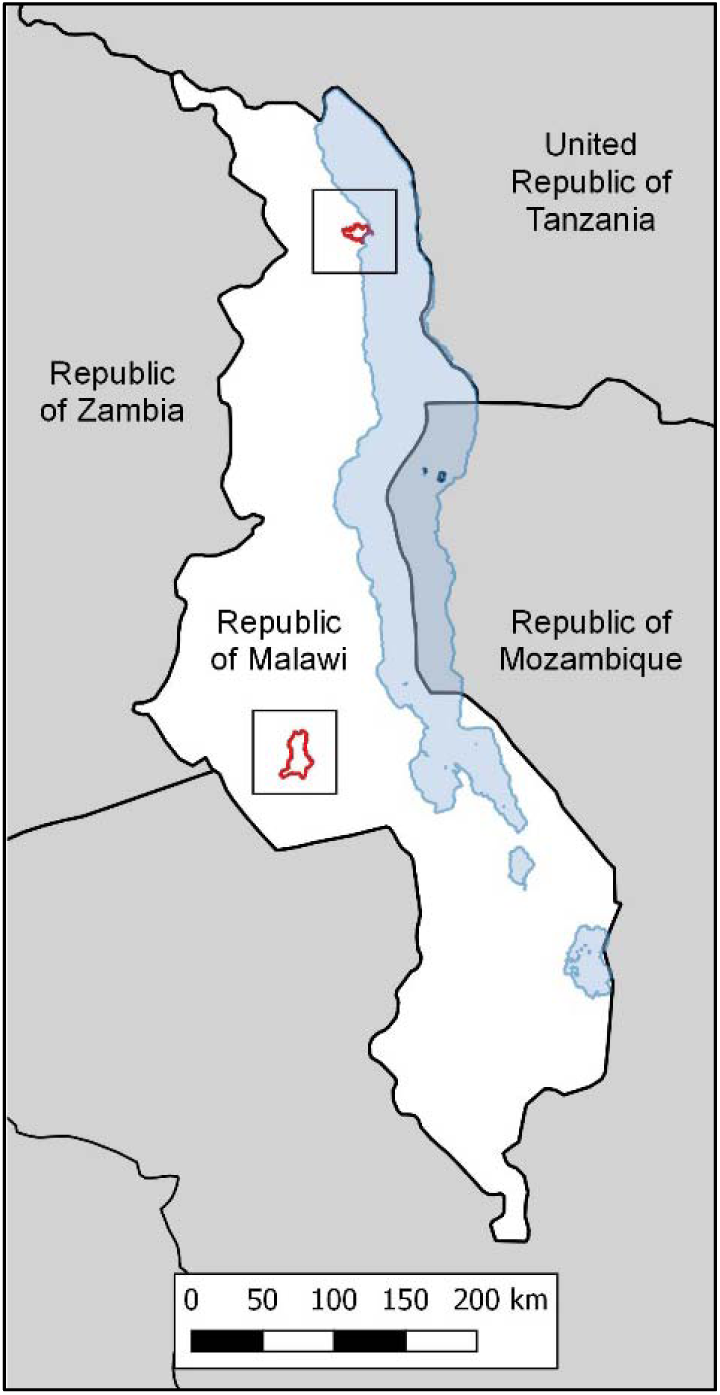
Study setting (national) Location of study sites within Malawi

**Figure 1B:**
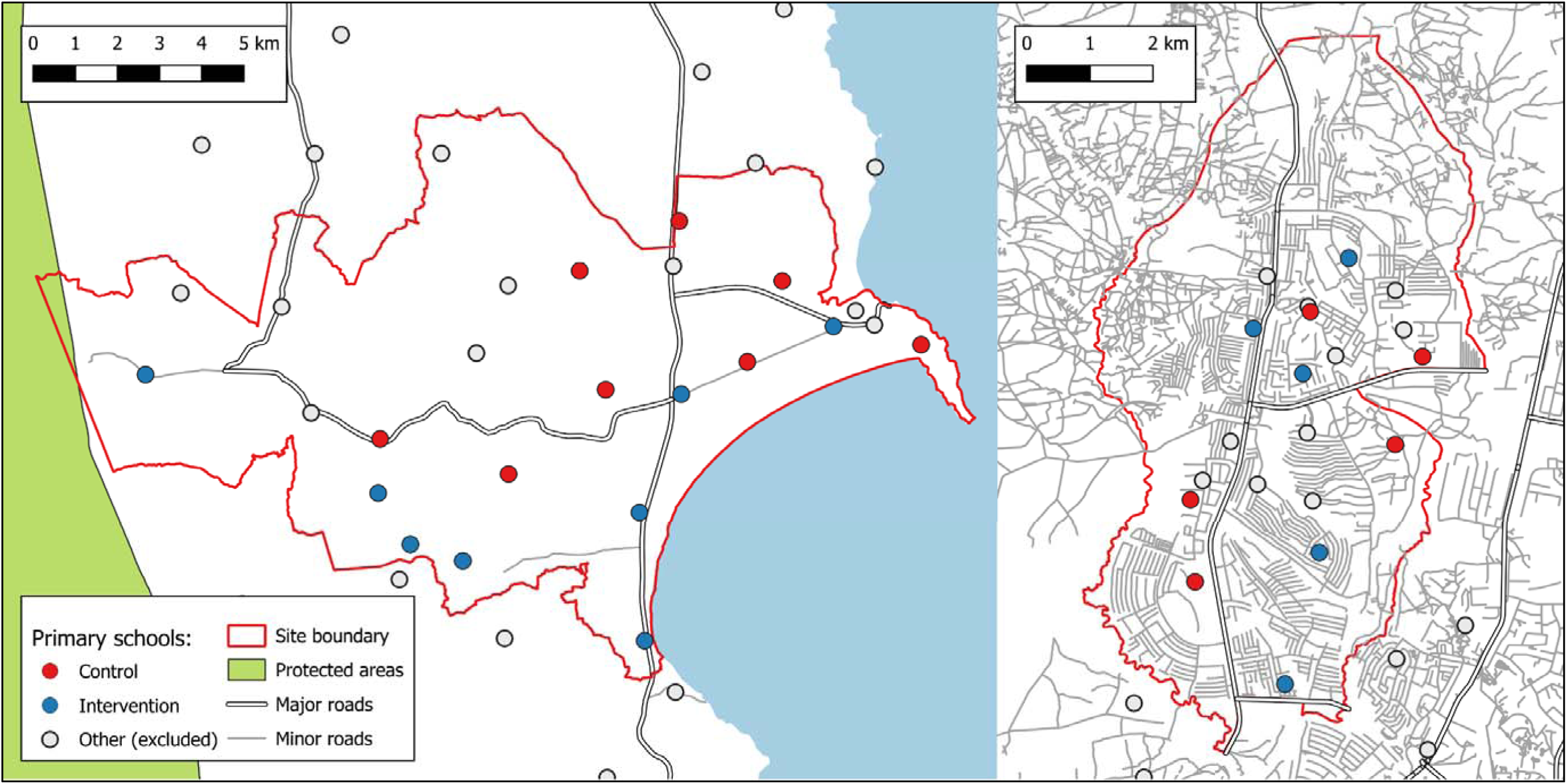
Study setting (sites) Map of study schools (clusters) within Karonga HDSS (left panel) and Lilongwe areas 25, 49 & 50 (right panel)

### 4.4. Intervention

The intervention content was adapted from a previously evaluated intervention in China [16] and pre-existing health education materials in Malawi. These materials were synthesised through co-creation workshops with stakeholders from health and education sectors, and refined following piloting [17]. The intervention had three main thematic areas: (i) physiological function and health consequences of excess salt, (ii) awareness and identification of sources of dietary salt (iii) strategies and solutions for reducing dietary salt intake; and was delivered through three complementary methods: classroom lessons, household learning, and participatory activities. Lessons were led by the regular standard 6 class teacher and consisted of didactic instruction and other routine teaching methods. Home-based learning to support the content of the lessons were delivered in the form of worksheets and involved predominantly inquiry-based methods (e.g. “identifying sources of salt in my home”). Participatory meetings included both school-based activities with adolescents (e.g. music and drama performances), and meetings with parents and caregivers facilitated by the class teacher to discuss salt reduction. In addition to a printed pupil and teachers handbook, schools were provided with illustrated posters to support the teaching materials. The intervention was delivered over a period of 10 weeks (1 school term) for approximately one hour per week. Immediately before the school term began, the class teacher received a three-day training in the programme delivered by staff from the MOHP and MOEST and their affiliates. Following this schools also received on-going support from the study team, who conducted spot-check observations of the intervention as it was being implemented.

### 4.5. Randomisation

A total of 26 school were eligible to participate in the study (Lilongwe=10, Karonga=16). Random assignment of study clusters was stratified by location: in Karonga as rural vs. semi-rural (12:4) and in Lilongwe as within Area 25 vs. outside Area 25 (5:5). Random assignment was conducted in two stages to permit balanced implementation across two school terms without revealing allocation to arm to schools in term 2 before completion of baseline assessments. Randomisation to term 1 or term 2 (1:1) was done within strata by an independent statistician, and randomisation to intervention or control (1:1) subsequently conducted at a public ceremony following completion of baseline measurements in all schools.

### 4.6. Participants

At each school, the enrolment register(s) for standard 6 were transcribed and used to conduct a physical rollcall by a study enumerator to confirm current enrolment. All adolescents potentially eligible to participate (aged 11-14 and currently enrolled) were randomly ranked and the first 30 adolescents issued with an invitation letter addressed to their parent or caregiver requesting a household visit to conduct the survey. Attempts were made to contact and enrol randomly selected adolescents who were absent up to three times.

At the household visit, adolescents were ineligible if the household planned to permanently leave within the next 12 months, an adolescent had already been sampled from that household, or the adolescent’s parent or caregiver did not reside within the same household. From each recruited adolescent, we invited their parents or caregivers for assessment (adult participant). If only one parent or caregiver was recruited, one other adult (>18 years) residing within the household was invited to participate, selected in order of closest age to the recruited parent or caregiver.

### 4.7. Outcomes

Co-primary outcomes were changes in adolescent urinary salt and SBP (3 months vs. baseline). Secondary outcomes were changes in adolescent DBP, adult urinary salt, and adult SBP and DBP (3 months vs. baseline), plus all outcomes at 12 months (vs. 3 months and baseline). Based on rapid literature review, pre-specified algorithms excluded likely incomplete urine samples. Adolescent urine samples were excluded if: (i) >1 self-reported missed void, (ii) 24h volume <300ml, or (iii) total creatinine <0·1 mmol/l. Adult urine samples were excluded if: (i) >1 self-reported missed void, (ii) 24h volume <500ml, or (iii) total creatinine <6 mmol/l (men) or <4 mmol/l (women). The mean of both height and weight measurements, and mean of the second and third BP readings, were calculated for analysis. Height-for-age and BMI-for-age z-scores in adolescents were calculated using WHO Child Growth Standards 2007 [18]. Age, sex, and height-adjusted blood pressure z-scores were calculated using reference standards by Flynn et al [19].

### 4.8. Procedures

Salt (sodium chloride) excretion was assessed by a single 24-hour urine collection. Participants were instructed by a trained enumerator and provided with equipment for urine collection both at home and to carry to school or work. The participant was instructed to collect all urine passed, and to continue even if a void was missed or spilt. After confirming the instructions, the participant was requested to empty their bladder and initiation time recorded. Shortly before completion of the 24-hour period, the enumerator returned to the household and requested the participant to pass any remaining urine before recording the time of completion and number of urine voids reported missed. To support adolescents to complete urine collection, schools were also requested to make a secure space available for adolescents to store equipment and specimens during school hours. Due to local restrictions on prolonged contact in response to the COVID-19 pandemic, no urine samples were collected at the 12-month survey. Urine specimens were measured for volume before being processed for sodium and creatinine concentrations using a Beckman Coulter AU480 analyser. Urine sodium was assayed via the crown ether membrane electrode method, and urine creatinine tested via the kinetic Jaffe method.

BP measurements were conducted after 30 minutes of rest and repeated three times at 5 minutes resting intervals. Measurements were collected on the right arm (or left if the participant had a condition that precluded measurement on the right arm) using a digital sphygmomanometer (OMRON M6 or equivalent) with appropriately sized cuff. Weight and height were measured twice following removal of shoes and outer clothing, using a weighing scale and stadiometer.

Compliance to the intervention, defined as the sampled adolescent having attended the lesson or activity, or completing the household activity worksheet, was assessed using attendance registers completed by the class teacher following completion of each intervention lesson or activity. Owing to the unblinded nature of the study and to supplement the parallel process evaluation, a post-implementation questionnaire was additionally conducted as part of the 12-month survey to explore the proportion of participants able to report the intervention focus (i.e. unblinding), and exposure to the intervention within the community (i.e. contamination).

### 4.9 Statistical analysis

Analysis of the primary and secondary outcomes were pre-specified and approved by an independent trial steering committee. Summary statistics were used to describe school (cluster) and individual characteristics at baseline. Differences in urinary salt and blood pressure between arms at 3 and 12-months respectively after baseline were estimated using mixed-effects models. The primary analysis model included a random intercept for school and adjusted for baseline, treatment, and trial stratification factors (Karonga or Lilongwe, school type, and term). All mixed-effect models were fitted using restricted maximum likelihood with the Kenward-Roger adjustment to the degrees of freedom. Separate level-1 (i.e. participant) residual variances were fitted for each arm. The primary analysis assumed missing outcomes were missing at random given baseline, treatment and the design variables. Data analysis was conducted using Stata 16.

Prespecified supplementary analyses were conducted to assess the effect of the intervention on urinary salt at 3-months amongst (1) adolescents only, after additionally adjusting for compliance to the intervention and (2) adolescent subgroups (by rural site, urban site, male sex, and female sex). We further conducted post-hoc supplementary analyses using formal tests of interaction to explore the effect of the intervention on urinary salt at 3-months amongst (1) participants with relatively high urinary salt at baseline, and (2) participants who reported the intervention focus on salt at the 12-month survey.

## 5. RESULTS

### 5.1. Recruitment and baseline characteristics

Table 1 describes the baseline characteristics of participants. A total of 1103 adolescents were sampled, with 732 adolescents subsequently completing the baseline assessments at a household visit where a further 1238 adults were also recruited (mean 1·7 adults recruited per adolescent). The characteristics of both adolescents and adults were balanced between study arms by all primary and secondary outcomes, and by individual characteristics of sex, age, weight, height, BMI and health status (previous diagnosis of hypertension or currently taking treatment for hypertension, adults only). Overall, mean urinary salt of participants (before applying any exclusion criteria) was 3·9g/day (SD=2·15g/day) for adolescents (n=730, 2 missing values) and 4·3g/day (SD=2·50g/day) for adults (n=1231, 7 missing values); with 26·6% of adolescents and 33·1% of adults therefore estimated to be consuming ≥5g salt/day. Mean systolic BP (SBP) of participants was 108·7mmHg (SD=11·78mmHg) for adolescents and 122.0mmHg (SD=0·5mmHg) for adults, with 16·6% of adolescents and 45·6% of adults having an SBP≥120mmHg (elevated BP); and 4·7% of adolescents and 25·4% of adults having an SBP≥130mmHg (high BP).

**Table 1:** Baseline school, adolescent, and parent characteristics by study group.

| Characteristic* | Control | Intervention |
| --- | --- | --- |
| **School characteristics:** | **N=13** | **N=13** |
| School location |  |  |
| Karonga HDSS, *n=* | 8 | 8 |
| Lilongwe (Area 25, 48 & 49) , *n=* | 5 | 5 |
| Academic term |  |  |
| Term 1 (October-December 2019), *n=* | 6 | 6 |
| Term 2 (January-March 2020), *n=* | 7 | 7 |
| School size |  |  |
| No. of children enrolled | 1895 (2480) | 1835 (1423) |
| Adolescents per school |  |  |
| No. of adolescents sampled, *Median (IQR)* | 43 (36-50) | 40 (34-50) |
| No. of adolescents assessed, *Median (IQR)* | 29 (27-30) | 28 (26-30) |
| **Adolescent characteristics** | **n=370** | **n=362** |
| Demographic |  |  |
| Sex (male), *n= (%)* | 168 (45.4%) | 158 (43.7%) |
| Age (years) | 12.9 (1.1) | 13.0 (1.0) |
| Anthropometry |  |  |
| Weight (kg) | 39.3 (8.5) | 39.1 (8.3) |
| Height (cm) | 145.9 (8.4) | 147.2 (8.6) |
| Height-for-age z-score | -1.3 (1.0) | -1.2 (1.2) |
| BMI-for-age z-score | -0.3 (1.0) | -0.4 (1.0) |
| Urine<sup>†</sup> |  |  |
| Valid urine sample, *n=* | 323 | 303 |
| Urinary salt (g/24h) | 4.3 (2.1) | 4.2 (2.2) |
| Urinary volume (ml/24h) | 815.7 (406.0) | 805.0 (368.4) |
| Blood pressure |  |  |
| Valid BP measurement, *n=* | 364 | 360 |
| Systolic BP (mmHg) | 108.6 (11.7) | 108.7 (11.9) |
| Diastolic BP (mmHg) | 68.1 (8.8) | 67.8 (8.8) |
| Systolic BP z-score | 0.5 (1.1) | 0.5 (1.1) |
| Diastolic BP z-score | 0.6 (0.9) | 0.6 (0.8) |
| **Adult characteristics** | **n=623** | **n=615** |
| Demographic: |  |  |
| Sex (male), *n=* | 250 (40.2%) | 244 (39.7%) |
| Age (years) | 41.2 (13.2) | 41.2 (12.3) |
| Anthropometry: |  |  |
| Weight (kg) | 63.8 (13.3) | 62.6 (12.9) |
| Height (cm) | 160.2 (8.0) | 160.0 (8.9) |
| BMI (kg/m<sup>2</sup>) | 24.9 (5.2) | 24.5 (4.9) |
| Urine<sup>‡</sup> |  |  |
| Valid urine sample, *n=* | 481 | 447 |
| Urinary salt (g/d) | 5.15 (2.72) | 5.07 (2.52) |
| Urinary volume (ml/d) | 1169 (560) | 1213 (627) |
| Blood pressure: |  |  |
| Valid BP measurement, *n=* | 611 | 612 |
| Systolic BP (mmHg) | 122.0 (16.7) | 122.0 (18.9) |
| Diastolic BP (mmHg) | 77.3 (10.89) | 77.9 (12.0) |
| Health status: |  |  |
| Previously diagnosed as hypertensive, *n= (%)* | 87 (14.0%) | 82 (13.3%) |
| Currently taking treatment for hypertension, *n= (%)* | 27 (4.3%) | 28 (4.6%) |

\*: Values are mean (SD) unless otherwise stated. †: For subset of adolescents meeting inclusion criteria that, at baseline and primary endpoint, all of: (a) no more than one self-report of missed urine void; (b) urine volume > 300 mL/24h; (c) total creatinine > 0.1 mmol/kg. ‡: For subset of adults meeting inclusion criteria that, at baseline
and primary endpoint, all of: (a) no more than one self-report missed void; (b) total urine < 500ml/24hl (c) total creatinine > 6mmol/l (men) or >4 mmol/l (women).

### 5.2. Compliance to intervention

Table 2 describes compliance to the intervention overall and by activity type, stratified by site and by term. There were no alterations to implementation of the intervention as it was designed, although school closures and other disruption caused by the response to the COVID-19 pandemic led to disruption to community activities in Lilongwe in term 2. Overall participation in the intervention was high, with 87·4% of intervention components either attended (if component was a lesson or community meeting) or completed (if component was a household activity) by the sampled adolescents allocated to the salt reduction programme. By component type, compliance was highest to the school lessons and lowest to the community activities; and moderately higher during term 1 compared to term 2; this pattern was consistent between sites.

**Table 2:** Summary of intervention compliance.

|  | Mean % of all intervention components completed (N*) | By intervention component type: |  |  |
| --- | --- | --- | --- | --- |
|  |  | Mean % of lessons attended (N*) | Mean % of household activities completed (N*) | Mean % of school meetings attended (N*) |
| Karonga (N=8 schools): |  |  |  |  |
| Term 1 | 90.5% (118) | 93.5% (118) | 96.3% (118) | 78.3% (118) |
| Term 2 | 88.6% (106) | 88.2% (106) | 91.1% (106) | 84.0% (106) |
| Lilongwe (N=5 schools): |  |  |  |  |
| Term 1 | 90.7% (50) | 96.1% (47) | 64.0% (50) | - |
| Term 2 | 77.9% (69) | 86.7% (55) | 87.0% (69) | 23.5% (55) |
| Overall (N=13 schools): |  |  |  |  |
| Total | 87.4% (343) | 91.0% (326) | 88.1% (343) | 69.6% (279) |

\*: Number of sampled adolescents for whom participation was recorded for at least one component of intervention or component group. Compliance defined as attendance to any one of 12 intervention components
where adolescent was eligible to participate (i.e. not left school or sick) and component was done. Attendance at school meetings could not be recorded during term 1 in Lilongwe.

### 5.3. Co-primary outcome

Table 3 describes the study co-primary outcome of urinary salt and SBP amongst adolescents at 3-month survey. There were 8 adolescent participants in each study arm lost to follow-up from baseline, with a total of 716 adolescent participants assessed at the 3-month survey immediately following completion of the intervention (Figure 2). After excluding adolescent participants who did not submit a urine sample or urine samples that did not meet the inclusion criteria, there were 434 (217 in each study arm) urine samples available for analysis.

**Figure 2:**
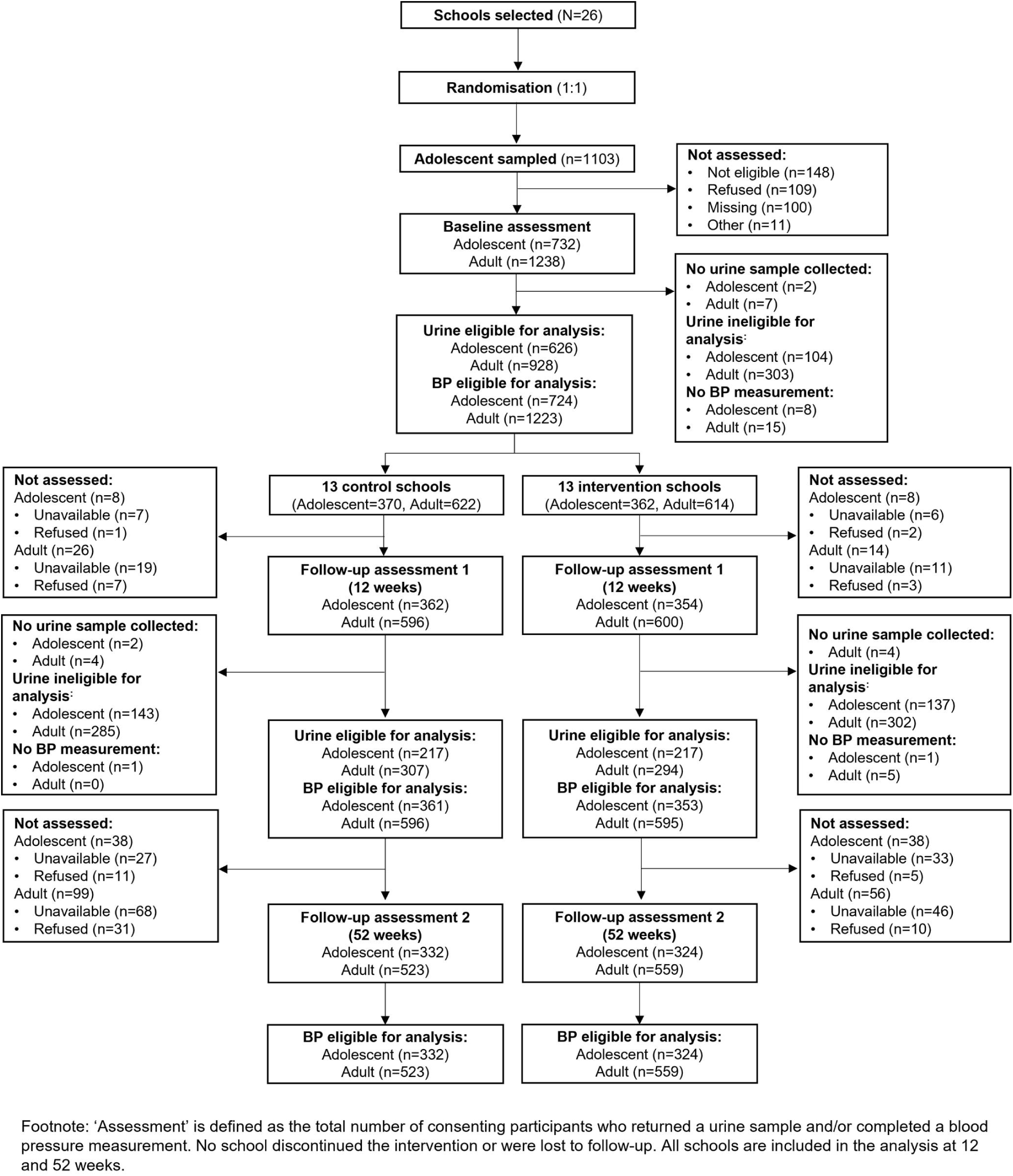
Trial profile.

**Table 3:**
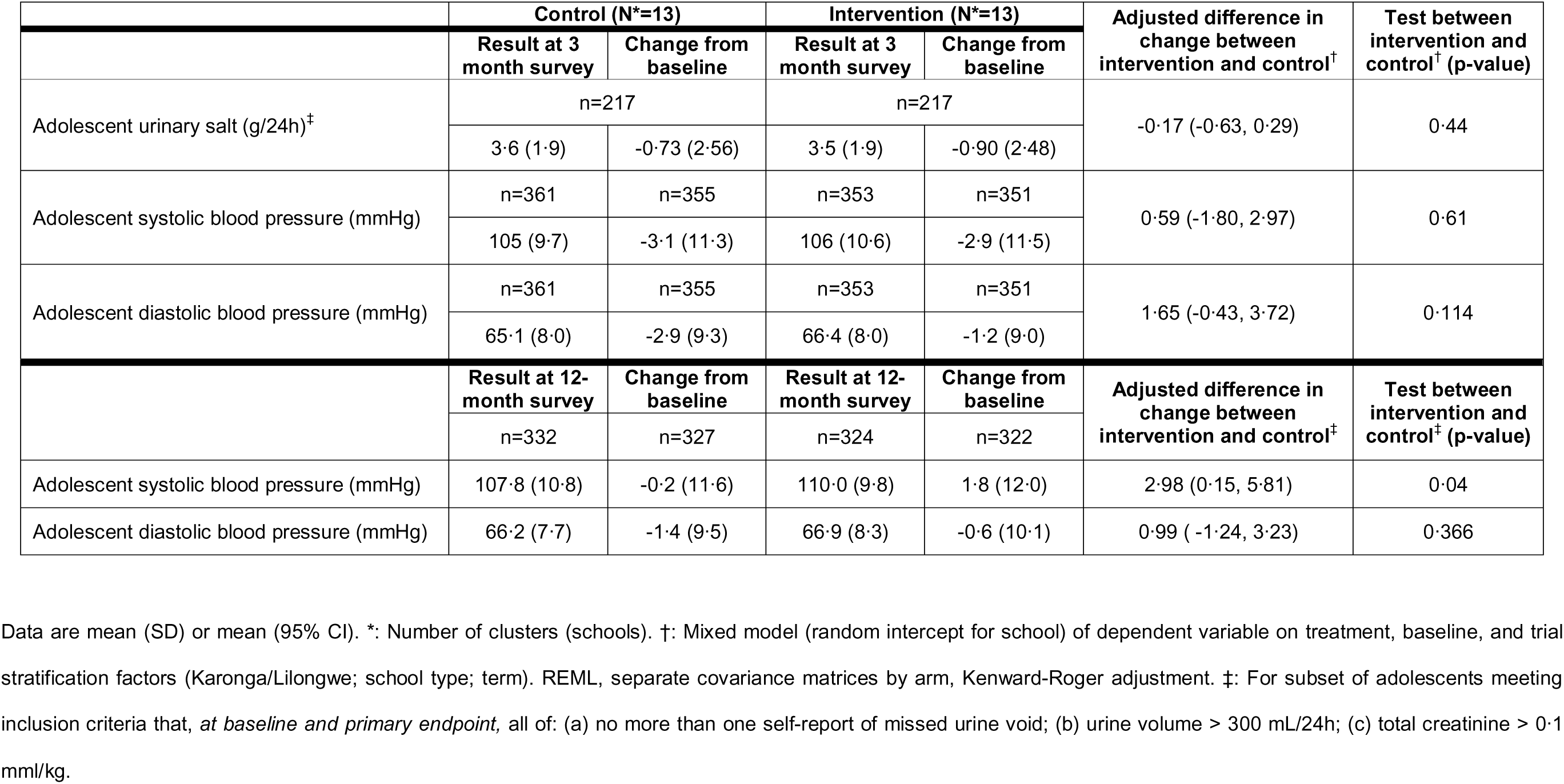
Co-primary outcome (effect of intervention on urinary salt and systolic blood pressure of adolescents at end of intervention (3-month survey)) and secondary outcomes amongst adolescents (effect of intervention on diastolic blood pressure at end of intervention, and systolic and diastolic blood pressure at end of study (12-month survey))

At 3-months, adolescents in the school-based salt reduction programme (intervention) had a mean reduction in urinary salt from baseline of -0·90g/day (SD=2·48); whereas adolescents in the routine curriculum (control) had a mean reduction in urinary salt from baseline of - 0·73g/day (SD=2·56). While adolescents urinary salt declined to a greater extent amongst adolescents in the salt reduction programme at 3-months; there was no significant difference in change in adolescent urinary salt between arms (-0·17g/day, 95% CI -0·63 to 0·29, p=0·44).

After excluding adolescent participants who did not have a valid SBP measurement at baseline or at 3 months, there were 706 BP measurements available for analysis at 3-months. Consistent with the outcome of urinary salt, there was also no significant difference in change in adolescent SBP between arms (-0·59mmHg, 95% CI: -1·80 to 2·97, p=0·61).

### 5.4. Secondary outcomes

Table 3 describes secondary outcomes amongst adolescents. Consistent with adolescent SBP, there was no significant different in change in adolescent DBP between study arms at 3-months (1·65mmHg, 95% CI: -0·43 to 3·72, p=0·114).

Loss to follow-up at 12-months was moderate amongst adolescents (n=76, 10·3%) and balanced between study arms. At 12-months, SBP was significantly higher amongst intervention-arm adolescents compared to control-arm adolescents (2·98mmHg, 95% CI 0·15 to 5·81, p=0·04), but there was no difference in change in adolescent DBP at 12-months (0·99mmHg, 95% CI -1·24 to 3·23, p=0·37).

Table 4 describes secondary outcomes, amongst adults. There were 26 adult participants in each study arm lost to follow-up from baseline, with a total of 1196 adult participants assessed at the 3-month survey immediately following completion of the intervention. After excluding invalid urine samples and participants who did not return a urine or participate in a BP measurement, there were 601 urine samples and 1191 BP measurements available for analysis at 3-months. There was no statistically significant difference in change between arms in adult urinary salt (-0·22g/day, 95% CI -0·59 to 0·16, p=0·26), adult SBP (0·81mmHg, 95% CI -0·99 to 2·61, p=0·36), or adult DBP (0·87mmHg, 95% CI -0·69 to 2·53, p=0·26) at 3-months.

**Table 4:** Secondary outcomes amongst adults: effect of intervention on (i) urinary salt and blood pressure at end of intervention (3-month survey) and (ii) blood pressure at end of study (12-month survey)

|  | Control (N*=13) |  | Intervention (N*=13) |  | Adjusted difference in change between intervention and control<sup>†</sup> | Test between intervention and control<sup>†</sup> (p-value) |
| --- | --- | --- | --- | --- | --- | --- |
|  | Result at 3-month survey | Change from baseline | Result at 3-month survey | Change from baseline |  |  |
| Adult urinary salt (g/24h)<sup>‡</sup> | n=307 |  | n=294 |  | -0.22 (-0.59, 0.16) | 0.261 |
|  | 4.25 (2.45) | -0.90 (3.07) | 4.09 (2.20) | -0.97 (3.00) |  |  |
| Adult systolic blood pressure (mmHg) | n=596 | n=585 | n=595 | n=593 | 0.81 (-0.99, 2.61) | 0.36 |
|  | 117.1 (15.3) | -4.76 (11.9) | 117.0 (16.7) | -4.47 (13.3) |  |  |
| Adult diastolic blood pressure (mmHg) | n=596 | n=585 | n=595 | n=593 | 0.87 (-0.69, 2.53) | 0.26 |
|  | 75.14 (10.57) | -2.13 (8.93) | 76.10 (11.24) | -1.53 (9.42) |  |  |
|  | Result at 12-month survey | Change from baseline | Result at 12-month survey | Change from baseline | Adjusted difference in change between intervention and control<sup>‡</sup> | Test between intervention and control<sup>‡</sup> (p-value) |
|  | n=523 | n=514 | n=559 | n=556 |  |  |
| Adult systolic blood pressure (mmHg) | 119.9 (16.2) | -1.85 (12.22) | 120.2 (18.25) | -1.77 (13.45) | 0.23 (-1.83, 2.29) | 0.82 |
| Adult diastolic blood pressure (mmHg) | 77.3 (9.82) | 0.16 (8.78) | 77.6 (12.9) | -0.44 (9.55) | -0.09 (-1.94, 1.77) | 0.92 |

Data are mean (SD) or mean (95% CI). \*: Number of clusters (schools). †: Mixed model (random intercept for school) of dependent variable on treatment, baseline, and trial stratification factors (Karonga/Lilongwe; school type; term). REML, separate covariance matrices by arm, Kenward-Roger adjustment. ‡: For subset of adults meeting inclusion criteria that, *at baseline and primary endpoint*, all of: (a) no more than one self-report missed void; (b) total urine < 500ml/24hl (c) total creatinine > 6mmol/l (men) or >4 mmol/l (women).

Loss to follow-up at 12-months was also moderate amongst adults (n=155, 12·1%) but was greater amongst control-arm adult participants (n=99, 15·9%). Consistent with results at 3-months, there was little difference in change in adult SBP (0·23mmHg, 95% CI -1·83 to 2·29, p=0.82) or adult DBP (-0·09, 95% CI -1·94 to 1·77, p=0·92) at 12-months.

### 5.5. Supplementary analyses

Prespecified supplementary analyses are reported in supplementary table 1. After adjusting for individual compliance to the intervention (Karonga only), there was no evidence of a dose-effect of the intervention on change in urinary salt amongst adolescents at 3-months (-1·38g/day, 95% CI -3·46 to 0·70, p=0·191). Analysis of adolescent urinary salt stratified by site (rural, urban) or sex (male, female) also showed no significant differences in intervention effect within these subgroups.

Post-hoc sub-group analyses exploring the effect of the intervention amongst participants with relatively high baseline urinary salt are reported in supplementary table 2. Amongst adults with baseline urinary salt of >4g/day, we observed a significant reduction in urinary salt at 3-months amongst intervention-arm adults as compared to control-arm adults (-0·57g/day, 95% CI -1·09 to 0·05, p=0·032). Similarly, amongst adolescents and adult urine samples with baseline salt >3·5g/24h (n=421), there was a significant reduction in urinary salt amongst intervention-arm participants at 3-months (-0·4g/day, 95% CI -0·67 to -0·13, p=0·056).

Finally, results of the post-trial questionnaire conducted at 12-months with adolescents are reported in supplementary table 3 and 5. Amongst control-arm adolescents, 13·6% (n=63) reported “salt” as the focus of the trial as compared to 54·8% (n=278) of intervention-arm adolescents; and 17·1% (n=57) of control-arm participants reported to have seen the intervention handbook. Post-hoc sub-group analysis demonstrated evidence of an interaction between awareness of the trial topic and change in adolescent urinary salt at 3-months (-0·80g/day, 95% CI -1·54 to -0·06) (supplementary table 7). However, when stratified by study arm, there was no evidence of an interaction between difference in change in adolescent urinary salt at 3-months and awareness of the trial topic in either control (0·65g/day, 95% CI -0·09 to 1·39) or intervention (-0·29g/day, 95% CI -0·84 to 0·26) arms (supplementary table 8). These results were broadly consistent amongst adults (supplementary table 4 and 6), with no evidence of an interaction between change in urinary salt and awareness of the trial topic overall (supplementary table 7) or by study arm (supplementary table 8).

## 6. DISCUSSION

In this cluster-randomised trial, a school-based programme to promote dietary salt reduction did not reduce the mean salt intake or blood pressure of adolescents or their parents and caregivers in Malawi. Despite high levels of participation, there was no difference in the urinary salt levels of adolescents immediately following the end of the intervention, or any pre-specified secondary outcome, when compared to those who continued to receive the routine school curriculum. This is the first trial of a school-based salt reduction intervention to be conducted in Africa, within a low-income country, or a rural setting. It provides an important contribution to the limited evidence base addressing salt intake and hypertension in low-income settings such as Malawi, evidence for school-health strategies in contributing more broadly to NCD policy, and necessary direction for future research.

Our results contrast with an earlier school-based salt reduction study in urban northern China, where high baseline salt intakes (7·3g/day in children, 12·6g/day in adults) were reduced by -1·9g/day and -2·9g/day respectively after three months [16]. However, subsequent evaluations of similar interventions in settings with lower baseline salt intake have shown mixed evidence for effectiveness. In south-east China, scale-up of a similar school-based salt reduction intervention demonstrated no effect on children, and more modest reduction in adults (-1·1g/day, baseline 9·0g/day) at 12 months [20]. Evaluation of an ‘app-based’ salt reduction intervention delivered through schools in China also did not reduce the salt intake of children, but achieved a modest reduction in adult intake at 12 months [21] that was not sustained at 24 months [22]. Elsewhere, a comparable school-based intervention evaluated in northern India demonstrated modest reductions in children’s salt intake (-0·5g/day), albeit assessed by dietary recall [23].

A large body of evidence demonstrates the effectiveness of behaviour change interventions to reduce adult salt intake by >1g/day [24]. For this study, we hypothesised that engaging adolescents as ‘agents of change’, supported by the direct involvement of teachers, would be an effective way of promoting salt reduction amongst parents and caregivers. This hypothesis was based on formative work conducted as part of the intervention development that identified adolescents interest in both their own and their caregivers health, parental value of school-health messages, and the involvement of adolescents in food preparation [17]. Similar interventions have previously been demonstrated in Africa as effective approaches for control of malaria [25] and promotion of sanitation [26]. In this study we observed no change in the salt intake of adults overall, although post-hoc analysis suggests that this mechanism may have been effective amongst adults with relatively high (>4g/day) baseline salt intake, suggesting that there may have been an effect amongst those at highest risk. Such a theory of change is likely to be highly contextual, and it is therefore unclear to what extent this pathway may have been ineffective at reducing the salt intake of adults overall for the same reasons as for adolescents, or whether the pathway itself - and the way in which it was delivered - was ineffective in this setting for adults with moderate salt intake.

A number of studies have demonstrated the role of schools in the prevention of NCDs more broadly including prevention of obesity [27] and promotion of physical activity [28], although predominantly in better resourced middle and high-income settings. While a major strength of school-health interventions is their efficiency in reaching a large number of children, they are limited by the amount of school time that can realistically be allocated to them. In this study, the intended ‘dose’ of the intervention was the maximum amount of time available in the routine school curriculum for health education (approximately 1 hour/week) over a single term with limited external supervision and previous teacher experience in delivery of health education, which may have been insufficient to effect behaviour change. A common feature of the education system in low-income settings is large teacher to pupil ratios, in addition to long-standing concerns about poor learning outcomes [29], which may also have limited the effectiveness of this intervention in this setting, compared to better-resourced settings.

The strengths of this study include the participation of communities in both rural and urban Malawi, with excellent recruitment and minimal loss to follow-up despite control measures introduced during the government response to the COVID-19 pandemic. The primary outcome of this study used 24-hour urine collection to estimate dietary salt, combined with rigorous inclusion criteria that included measurement of urinary creatinine. This study was well-powered, with a baseline-adjusted mean salt intake SD=1·85 (compared to a predicted SD=2·5), and ICC of 0·15 (compared to a predicted ICC=0·05), giving >80% power to detect an intervention effect of 0·9g/day. Finally, the intervention itself, which was developed through a thorough process of co-creation and piloting with stakeholders from both the education and health sectors in Malawi, was implemented by the routine education service and had high levels of participation and acceptability [17].

This study has several limitations. Given the nature of the intervention and study procedures it was not possible to fully blind participants to the outcome of urinary salt, or to maintain non-contamination of control-arm participants, a challenge that has previously been recognised for randomised trials of educational interventions [30]. Our 12-month survey identified knowledge amongst both control and intervention arm participants as to the focus of the intervention, and exposure to the intervention amongst control-arm participants. While there was no evidence of an interaction between these factors and change in urinary salt when stratified by study arm, we cannot exclude the risk that this may have biased our results or diluted internal validity. In the absence of any standardised inclusion/exclusion criteria for 24-hour urine samples, we developed rigorous criteria that were based on a review of evidence and previous comparable studies. However, much of this evidence either lacks empirical data (e.g. minimum urine volume) or is reliant on evidence from clinical studies conducted outside SSA (e.g. minimum urinary creatinine), and therefore our criteria may not have been optimal for this study. Our criteria also resulted in a non-trivial proportion of urine samples being excluded, and as it was not feasible for us to re-assess participants whose urine was not eligible for analysis, reduced the sample size and statistical precision of our estimates. At baseline, salt intakes in our study population were moderate, with mean consumption below 5g/day in both adolescents and adults, potentially limiting the scope for measurable reductions over the (relatively) short intervention period.

## 7. CONCLUSION

In this randomised controlled trial conducted across rural and urban Malawi, we found no effect of a school-based salt reduction programme on the salt intake or blood pressure of adolescents or their parents and caregivers after one school term (3 months), compared to those who received the routine school curriculum (control). With high compliance and fidelity of delivery, this trial demonstrated that delivery of a school-based salt reduction intervention was feasible and acceptable within the education system in Malawi, but may have been limited by the short intervention period and modest baseline dietary salt intake. Schools remain a pragmatic platform for addressing the growing burden of NCDs more broadly amongst children and their families. Future research should focus on the feasibility and effectiveness of sustained school-based interventions targeting NCDs, and understanding how best these interventions can contribute to comprehensive population-level NCD prevention strategies.

## 9. ADDENDUM

## 9.1 Acknowledgments

We sincerely thank all those who participated in this trial, with special recognition of the residents and standard 6 school teachers of Karonga HDSS and Lilongwe areas 25, 48 & 49 who generously contributed to implementation and evaluation.

At the Malawi Epidemiology and Intervention Research Unit, we thank the team of enumerators, clinicians and drivers who contributed to data collection; in addition to those who contributed to the coordination, data management, and laboratory analysis of this trial (Esmie Banda, Louis Banda, Odala Chithodwe, Mabvuto Kamulayike, Cynthia Katundu, Justice Khosa, Marriot Kondowe, Alick Mwalwanda, Oddie Mwiba, Hazel Namadingo, Lawrence Nkhwazi, Laurence Tembo).

We also thank those at the University of Glasgow (Chris Chapman, Eleanor Grieve), LSHTM (Abena Amoah, Estelle McLean, Frankie Liew), and Ministry of Health & Population in Malawi (Beatrice Mwagomba) who contributed to the design, coordination and supervision of this trial.

## 9.4. Data availability statement

Data will be made available on publication through the LSHTM Data Compass (http://datacompass.lshtm.ac.uk). Requests for release of the data will be reviewed by the relevant institutional review boards.

## 9.5. Author contributions

ACC, EM, FM, JRG, JM, JRC and MKe contributed to conceptualisation, funding acquisition and methodology. AS, CM, MC, MKa, NP, SM and SWM contributed to investigation. ACC, DDK, EM, FM, JM, JRG, MKe contributed to supervision. SWM contributed to data curation and software. JRC, RB and SWM conducted the formal analysis. JRC, MC and SWM wrote the original draft of the manuscript. All authors reviewed and approved the final manuscript.

