## Supplementary Materials for "School-based programme to reduce dietary salt intake and blood pressure amongst adolescents and their parents: a cluster-randomised controlled trial in rural and urban Malawi"

**Supplementary table 1: Prespecified supplementary analysis – effect of intervention on urinary salt at end of intervention (3-months) amongst adolescent participants (1) adjusted for compliance to intervention and (2) sub-group analysis by trial site (rural, urban) and sex (male, female)**

|  | **Control (N*=13)** | | **Intervention (N*=13)** | | **Adjusted difference in change between intervention and control**^†^ | **Test between**  **intervention and**  **control**^†^ **(p-value)** |
| --- | --- | --- | --- | --- | --- | --- |
|  | **Result at 3-month survey** | **Change from baseline** | **Result at 3-month survey** | **Change from baseline** |  |  |
| Urinary salt (g/24h): adolescents only, adjusted for compliance^‡^ | n=153 | n=153 | n=141 | n=141 | -1·38 (-3·46 to 0·70) | 0·191 |
|  | 3·3 (1·82) | -0·8 (2·59) | 3·3 (1·91) | -0·8 (2·45) |  |  |
| Urinary salt (g/24h): adolescents only (rural site, Karonga) | n=153 | n=153 | n=141 | n=141 | -0·20 (-0·890, 0·497) | 0·551 |
|  | 3·3 (1·82) | -0·8 (2·59) | 3·3 (1·91) | -0·8 (2·45) |  |  |
| Urinary salt (g/24h): adolescents only (urban site, Lilongwe) | n=64 | n=64 | n=76 | n=76 | -0·51 (-1·14, 0·11)^∆^ | 0·108^∆^ |
|  | 4·3 (1·98) | -0·68 (2·48) | 3·8 (1·83) | -1·1 (2·55) |  |  |
| Urinary salt (g/24h): adolescents only (boys) | n=108 | n=108 | n=96 | n=96 | <0·01 (-0·58, 0·58) | 0·998 |
|  | 3·3 (1·62) | -0·72 (2·47) | 3·3 (1·75) | -0·58 (2·20) |  |  |
| Urinary salt (g/24h): adolescents only (girls) | n=109 | n=109 | n=121 | n=121 | -0·36 (-1·07, 0·35) | 0·305 |
|  | 3·9 (2·14) | -0·74 (2·64) | 3·6 (2·01) | -1·16 (2·67) |  |  |

Data are mean (SD) or mean (95% CI). *: Number of clusters (schools). †: Mixed model (random intercept for school) of dependent variable on treatment, baseline, and trial stratification factors (Karonga/Lilongwe; school type; term). REML, separate residual variance by arm, Kenward-Roger adjustment. ‡: (integer 0 – 10; available from Karonga only). ∆: Random effect model estimated school SD as 0, so model reported is standard linear regression. Unusually, regression sandwich variance estimator at school level gives effect of -0·51 (-0·95, -0·07), p=0·027, *i.e.* a smaller standard error, probably due to the relatively small number of schools (n=10) with between 6 and 21 students from each. §: For subset of adolescents meeting inclusion criteria that, *at baseline and primary endpoint,* all of: (a) no more than one self-report of missed urine void; (b) urine volume > 300 mL/24h; (c) total creatinine > 0.1 mml/kg. ¶: For subset of adults meeting inclusion criteria that, *at baseline and primary endpoint,* all of: (a) no more than one self-report missed void; (b) total urine < 500ml/24hl (c) total creatinine > 6mmol/l (men) or >4 mmol/l (women).

**Supplementary table 2: Post-hoc sub-group analysis 1 – effect of intervention on urinary salt at end of intervention (3-months) amongst (i) all participants (ii) all participants with baseline urinary salt >3.5g/24h and (iii) adult participants with baseline urinary salt >4g/24h**

|  | **Control (N*=13)** | | **Intervention (N*=13)** | | **Adjusted difference in change between intervention and control**^†^ | **Test between**  **intervention and**  **control**^†^ **(p-value)** |
| --- | --- | --- | --- | --- | --- | --- |
|  | **Result at 3-month survey** | **Change from baseline** | **Result at 3-month survey** | **Change from baseline** |  |  |
| Urinary salt (g/24h): all adolescent and adults ^§¶^ | n=524 | | n=511 | | -0·21 (-0·48 to 0·06) | 0·129 |
|  | 4·0 (2·27) | -0·8 (2·9) | 3·8 (2·10) | -0·9 (2·8) |  |  |
| Urinary salt (g/24h): adolescent and adults with baseline >3·5g/24h^§¶^ | n=219 | | n=204 | | -0·4 (-0·67 to -0·13) | 0·056^#^ |
|  | 4·6 (2·57) | -1·6 (3·15) | 4·32 (2·24) | -1·8 (2·97) |  |  |
| Urinary salt (g/24h): adults only with baseline >4g/24h^¶^ | n=187 | | n=177 | | -0·57 (-1·09 to -0·05) | 0·032^#^ |
|  | 4·7 (2·63) | -1·9 (3·18) | 4·3 (2·30) | -2·2 (2·91) |  |  |

Data are mean (SD) or mean (95% CI). *: Number of clusters (schools). †: Mixed model (random intercept for school) of dependent variable on treatment, baseline, and trial stratification factors (Karonga/Lilongwe; school type; term). REML, separate covariance matrices by arm, Kenward-Roger adjustment. ‡: (integer 0 – 10; Karonga only). §: For subset of adolescents meeting inclusion criteria that, *at baseline and primary endpoint,* all of: (a) no more than one self-report of missed urine void; (b) urine volume > 300 mL/24h; (c) total creatinine >0·1 mml/kg. ¶: For subset of adults meeting inclusion criteria that, *at baseline and primary endpoint,* all of: (a) no more than one self-report missed void; (b) total urine < 500ml/24hl (c) total creatinine > 6mmol/l (men) or >4 mmol/l (women). #: Models exclude random intercept for school as this was not significant (p=0·37 and p=0·60 respectively).

**Supplementary table 3: post-trial survey – adolescent participants, stratified by arm**

|  | **Control** | **Intervention** |
| --- | --- | --- |
|  | n=336 (51·1%) | n=322 (48·9%) |
| *Do you remember that you participated in a study on diet?* | | |
| No | 16 (4·8%) | 9 (2·8%) |
| Yes | 319 (94·9%) | 312 (96·9%) |
| Don't know | 1 (0·3%) | 1 (0·3%) |
| *Did you ever hear people talking about this study in the community?* | | |
| No | 219 (65·2%) | 156 (48·4%) |
| Yes | 117 (34·8%) | 166 (51·6%) |
| *Is salt mentioned as a main or key-topic* | | |
| Salt related | 59 (17·6%) | 196 (60·9%) |
| Health related | 184 (54·8%) | 85 (26·4%) |
| Don't know/other unrelated | 93 (27·7%) | 41 (12·7%) |
| *Have you seen any of these materials over the last year?* | | |
| No | 276 (82·9%) | 0 (·%) |
| Yes | 57 (17·1%) | 0 (·%) |
| *Have you changed any eating or drinking habits at home since last year?* | | |
| No | 244 (72·6%) | 124 (38·5%) |
| Yes | 92 (27·4%) | 198 (61·5%) |

**Supplementary table 4: post-trial survey – adult participants, stratified by arm**

|  | **Control** | **Intervention** |
| --- | --- | --- |
|  | n=462 (47·7%) | n=507 (52·3%) |
| *Do you remember that you participated in a study on diet?* | | |
| No | 8 (1·7%) | 9 (1·8%) |
| Yes | 453 (98·1%) | 497 (98·0%) |
| Don't know | 1 (0·2%) | 1 (0·2%) |
| *Did you ever hear people talking about this study in the community?* | | |
| No | 253 (54·8%) | 220 (43·4%) |
| Yes | 207 (44·8%) | 285 (56·2%) |
| Don't know | 2 (0·4%) | 2 (0·4%) |
| *Is salt mentioned as a main or key-topic* | | |
| Salt related | 63 (13·6%) | 278 (54·8%) |
| Health related | 329 (71·2%) | 189 (37·3%) |
| Don't know/other unrelated | 70 (15·2%) | 40 (7·9%) |
| *Have you seen any of these materials over the last year?* | | |
| No | 416 (90·6%) | 1 (100·0%) |
| Yes | 42 (9·2%) | 0 (0·0%) |
| Don't know | 1 (0·2%) | 0 (0·0%) |
| *Have you changed any eating or drinking habits at home since last year?* | | |
| No | 351 (76·0%) | 203 (40·0%) |
| Yes | 111 (24·0%) | 302 (59·6%) |
| Don't know | 0 (0·0%) | 2 (0·4%) |

**Supplementary table 5: post-trial survey – adolescent participants, stratified by site and arm**

|  | **Karonga** | | **Lilongwe** | |
| --- | --- | --- | --- | --- |
|  | **Control** | **Intervention** | **Control** | **Intervention** |
|  | n=219 (33·3%) | n=211 (32·1%) | n=117 (17·8%) | n=111 (16·9%) |
| *Do you remember that you participated in a study on diet?* | | |  |  |
| No | 12 (5·5%) | 9 (4·3%) | 4 (3·4%) | 0 (0·0%) |
| Yes | 207 (94·5%) | 201 (95·3%) | 112 (95·7%) | 111 (100·0%) |
| Don't know | 0 (0·0%) | 1 (0·5%) | 1 (0·9%) | 0 (0·0%) |
| *Did you ever hear people talking about this study in the community?* | | |  |  |
| No | 112 (51·1%) | 86 (40·8%) | 107 (91·5%) | 70 (63·1%) |
| Yes | 107 (48·9%) | 125 (59·2%) | 10 (8·5%) | 41 (36·9%) |
| *Is salt mentioned as a main or key-topic* | | |  |  |
| Salt related | 12 (5·5%) | 101 (47·9%) | 47 (40·2%) | 95 (85·6%) |
| Health related | 127 (58·0%) | 73 (34·6%) | 57 (48·7%) | 12 (10·8%) |
| Don't know/other unrelated | 80 (36·5%) | 37 (17·5%) | 13 (11·1%) | 4 (3·6%) |
| *Have you seen any of these materials over the last year?* | | |  |  |
| No | 190 (86·8%) | 0 (·%) | 86 (75·4%) | 0 (·%) |
| Yes | 29 (13·2%) | 0 (·%) | 28 (24·6%) | 0 (·%) |
| *Have you changed any eating or drinking habits at home since last year?* | | |  |  |
| No | 166 (75·8%) | 102 (48·3%) | 78 (66·7%) | 22 (19·8%) |
| Yes | 53 (24·2%) | 109 (51·7%) | 39 (33·3%) | 89 (80·2%) |

**Supplementary table 6: post-trial survey – adult participants, stratified by site and arm**

|  | **Karonga** | | **Lilongwe** | |
| --- | --- | --- | --- | --- |
|  | **Control** | **Intervention** | **Control** | **Intervention** |
|  | n=336 (34·7%) | n=344 (35·5%) | n=126 (13·0%) | n=163 (16·8%) |
| *Do you remember that you participated in a study on diet?* | | |  |  |
| No | 6 (1·8%) | 9 (2·6%) | 2 (1·6%) | 0 (0·0%) |
| Yes | 329 (97·9%) | 334 (97·1%) | 124 (98·4%) | 163 (100·0%) |
| Don't know | 1 (0·3%) | 1 (0·3%) | 0 (0·0%) | 0 (0·0%) |
| *Did you ever hear people talking about this study in the community?* | | |  |  |
| No | 138 (41·1%) | 97 (28·2%) | 115 (91·3%) | 123 (75·5%) |
| Yes | 196 (58·3%) | 246 (71·5%) | 11 (8·7%) | 39 (23·9%) |
| Don't know | 2 (0·6%) | 1 (0·3%) | 0 (0·0%) | 1 (0·6%) |
| *Is salt mentioned as a main or key-topic* | | |  |  |
| Salt related | 11 (3·3%) | 145 (42·2%) | 52 (41·3%) | 133 (81·6%) |
| Health related | 262 (78·0%) | 167 (48·5%) | 67 (53·2%) | 22 (13·5%) |
| Don't know/other unrelated | 63 (18·8%) | 32 (9·3%) | 7 (5·6%) | 8 (4·9%) |
| *Have you seen any of these materials over the last year?* | | |  |  |
| No | 315 (94·0%) | 0 (·%) | 101 (81·5%) | 1 (100·0%) |
| Yes | 19 (5·7%) | 0 (·%) | 23 (18·5%) | 0 (0·0%) |
| Don't know | 1 (0·3%) | 0 (·%) | 0 (0·0%) | 0 (0·0%) |
| *Have you changed any eating or drinking habits at home since last year?* | | |  |  |
| No | 266 (79·2%) | 159 (46·2%) | 85 (67·5%) | 44 (27·0%) |
| Yes | 70 (20·8%) | 183 (53·2%) | 41 (32·5%) | 119 (73·0%) |
| Don't know | 0 (0·0%) | 2 (0·6%) | 0 (0·0%) | 0 (0·0%) |

**Supplementary table 7: Post-hoc sub-group analysis 2 – effect of intervention on urinary salt and diastolic blood pressure at 3-months amongst adolescent and adult participants, stratified by reported recall about trial at 12-months**

|  | **Change from baseline** | | **Interaction test** |
| --- | --- | --- | --- |
|  | **“No”*** | **“Yes”*** |  |
| Adolescent urinary salt (g/24h)^†^ | 0·13  (-0·42, 0·68) | -0·80  (-1·54, -0·06) | p=0·038 |
| Adolescent diastolic blood pressure (mmHg) | 1·59  (-0·70, 3·88) | 2·55  (-0·35, 5·26) | p=0·54 |
| Adult urinary salt (g/24h)^†^ | 0·10  (-0·39, 0·59) | -0·72  (-1·65, 0·22) | p=0·134 |
| Adult diastolic blood pressure (mmHg) | 0·81  (-0·94, 2·57) | 0·24  (-2·34, 2·82) | p=0·68 |

* Mentioned “salt” in response to question “*Is salt mentioned as a main or key-topic?*”. † Among participants whose urine data met the criteria for inclusion in the primary analysis.

**Supplementary table 8: Post-hoc sub-group analysis 3 – effect of intervention on urinary salt and diastolic blood pressure at 3-months amongst adolescent and adult participants, stratified by reported recall about trial at 12-months and by trial arm**

|  | **Trial**  **arm** | **Change from baseline** | | **Adjusted difference in change**  **(“Yes”-“No”)**^†^ | **Test between**  **“Yes” and “No” (p-value)** ^†^ |
| --- | --- | --- | --- | --- | --- |
|  |  | **“No”*** | **“Yes”*** |  |  |
| Adolescent urinary salt (g/24h)^‡^ | Control | n=182 | n=35 | 0·650  (-0·092, 1·392) | 0·086 |
|  |  | -0·79 | -0·42 |  |  |
|  | Intervention | n=101 | n=116 | -0·291  (-0·837, 0·255) | 0·294 |
|  |  | -0·75 | -1·03 |  |  |
| Adolescent diastolic blood pressure (mmHg) | Control | n=298 | n=57 | -0·664  (-3·015, 1·686) | 0·579 |
|  |  | -2·69 | -4·16 |  |  |
|  | Intervention | n=157 | n=194 | -0·476  (-2·094, 1·141) | 0·563 |
|  |  | -1·91 | -0·70 |  |  |
| Adult urinary salt (g/24h)^‡^ | Control | n=278 | n=29 | 0·309  (-0·756, 1·374) | 0·568 |
|  |  | -0·92 | -0·74 |  |  |
|  | Intervention | n=138 | n=158 | -0·470  (-1·021, 0·081) | 0·094 |
|  |  | -0·65 | -1·25 |  |  |
| Adult diastolic blood pressure (mmHg) | Control | n=520 | n=65 | 0·938  (-1·441, 3·318) | 0·439 |
|  |  | -2·17 | -1·77 |  |  |
|  | Intervention | n=295 | n=298 | 0·325  (-1·142, 1·791) | 0·664 |
|  |  | -1·58 | -1·47 |  |  |

*Mentioned “salt” in response to question “*Is salt mentioned as a main or key-topic?”* †: Adjusted for same covariates as primary analysis (i.e. baseline and design variables). ‡: Among participants whose urine data met the criteria for inclusion in the primary analysis.

**Supplementary figure 1: Study timeline**

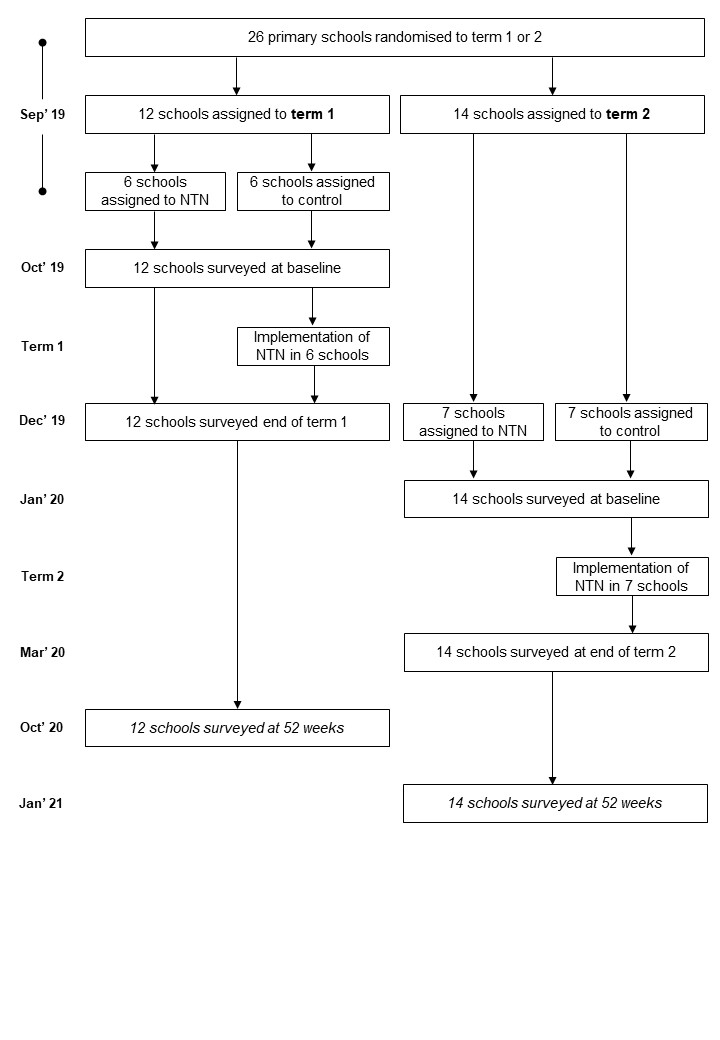

Caption: Timeline of study implementation. Note: In response to the COVID-19 pandemic, primary schools in Malawi were closed between late March–mid October 2020 and late January–early March 2021.
